# Age at Onset and Liability to Disorder: Estimating Covariances in Censored Populations

**DOI:** 10.64898/2026.08.04.26359043

**Authors:** Michael C. Neale, Hermine H. Maes, Lanie K. Mullins, Madhurbain Singh, Jared Balbona, Robert M. Kirkpatrick, Timothy R. Brick, Michael D. Hunter, Steven M. Boker, Luis Castro-de-Araujo, Andrew J. Schork, Morten D. Krebs, Joel A. Mefford

## Abstract

Studies of resemblance for disorders and other traits measured at the binary (yes/no) level between relatives frequently contain individuals who are currently in the negative category but who will become positive in future. For example, a 10-year-old may develop depression in the future, but is as yet unaffected. Such censoring can substantially bias estimates of correlation between relatives. To overcome this problem we develop a model for the association between liability to a disorder, and its age at onset. The model is designed for data from pairs of relatives to enable estimation of the correlation between an individuals’ liability to disorder and their age at onset. Usually, such information is not available at the individual level, because age at onset is uniquely available when onset has occurred. Lacking variation in disorder status, data from non-related persons cannot estimate the covariance between liability and age at onset. Data from relatives can resolve this issue when there is a correlation in liability between the relatives, because different age at onset distributions would be expected in concordant vs. discordant pairs of relatives. Greater severity and worse outcomes are often observed among those with earlier onset, so a correlation between disorder liability and age at onset seems likely in many cases. In this article we present the basic theory of the model, implemented as a mixture distribution, and an application to cannabis use in a Virginia Twin Study of Adolescent Behavioral Development. A negative association of (-.212) between age at onset an liability was found, with confidence intervals of −.263 to −.152, which do not cross zero. The method contrasts with Cox Proportional Hazards, in which disorder liability and onset timing are treated as a single dimension.

## Introduction

In behavioral genetic studies, it is common practice to assess correlations between pairs of relatives to see if they covary with degree of genetic similarity. The many twin studies published over the past half century are examples of this activity. The methodology has been used for continuous, ordinal and binary traits, and has been applied to many physical, psychiatric, and substance use disorders. A potential limitation of the application to these disorders is that unaffected individuals may not have reached the age at which their onset would occur. Some of these unaffected individuals will become affected in the future, whereas those whose disorder liability is too low will never contract the disorder Neale, Eaves, Hewitt, et al., 1989.

The literature on behavior genetics contains many examples of twin and family studies of populations that have not passed their age at risk. Previous analyses of psychiatric disorders and substance use (e.g., Kendler et al., 1992, 1995, 1999) are examples from our institute. These articles may have reported estimates that were biased due to censored observations of future first onsets of disorders. The degree of bias depends on the proportion of the sample still at risk of contracting the disorder, which will vary between disorders and between the populations under study – potentially hampering replication efforts. It is important to understand the effects of censoring on correlations between relatives. Ideally, analytical methods should be developed that will remove bias due to censoring: that is one goal of this article.

### Theory

In a sample where some individuals have yet to reach their age at onset, there are two possible states of those who are unaffected. They may be immune, which we might model as being below a threshold on a normal distribution of liability. In addition, it is possible that their age at onset has yet to occur. For a single individual, the likelihood of being affected would be:

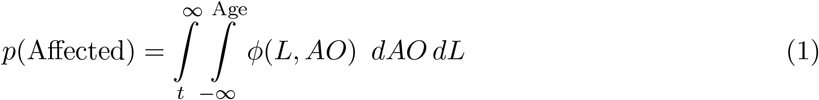

where: *ϕ* represents the multivariate normal probability density function, AO is the onset distribution and L is the liability to disorder distribution. In what follows, we assume that the age-at-onset (AO) distribution and the liability (L) distributions follow the bivariate normal distribution or the multivariate normal across relatives. This model for correlated liabilities (within-individual) is shown in the top left panel of Figure 1. Equation 1 applies when the actual age at onset is unknown - it is simply less than the age at assessment (Age). The likelihood of being affected, above the disorder threshold and older than the age of onset, is shown in the lower right panel of Figure 1. When ages at onset have been observed, the expression reduces to:

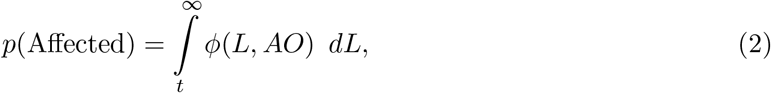

i.e., the integral over the bivariate liability and onset dimensions, with the age at onset held constant at AO. It is this situation, age at onset known for all affected individuals, that is the primary focus of this article. In part, this is because the statistical power in the absence of measured ages at onset seems likely to be very low, requiring very large sample sizes for accurate parameter estimates.

**Figure 1.**
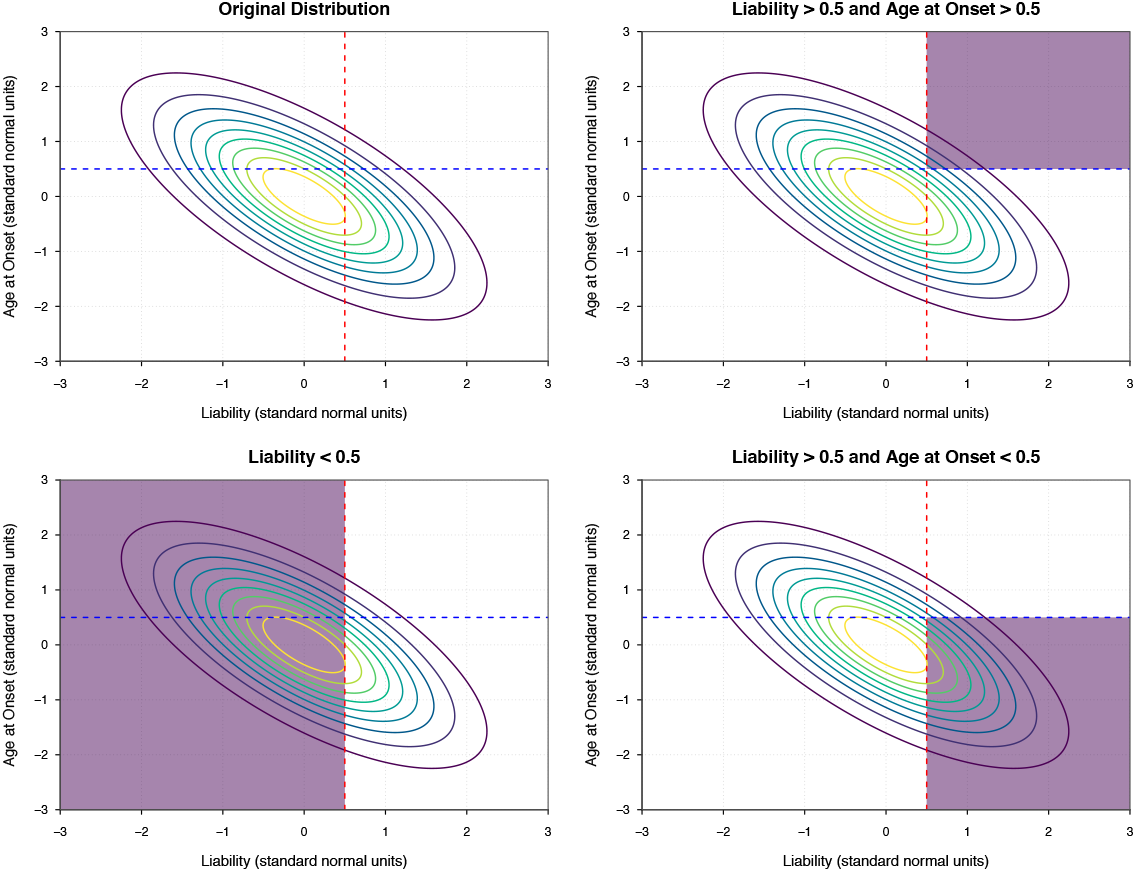
Bivariate normal contour plots depicting −.7 correlation between age at onset liability and disorder. Anticlockwise from top left: i) full population, ii) shaded region depicts individuals below liability threshold, iii) affected individuals above liability threshold and onset earlier than current age, and iv) individuals at risk but younger than their age at onset.

If an individual is unaffected, trivially we can note that *p*(Unaffected) = 1 *− p*(Affected). However, as noted above, this probability features a mixture of two types: i) those whose liability is below the threshold (for whom age at onset is irrelevant) and ii) those who are above the threshold but have yet to reach their age at onset. Thus the probability of being unaffected at a given age at assessment (“Age”) is

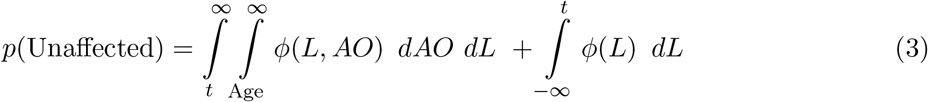

To estimate the covariance between liability to disorder, and age at onset is not possible with data from unrelated persons alone. This problem occurs because with a binary measure of disorder, only those affected have an age at onset. Since there is no variation in disorder status, it is not possible to directly measure its covariance. However, as has long been recognized in behavior genetic studies, data from relatives can provide valuable new information. First, if relatives correlate positively for liability to disorder, then the average liability of concordant pairs will be higher than that of discordant pairs. Second, if liability is a cause of age at onset (e.g., high liability associates with earlier age at onset), then the average age at onset of concordant pairs would be expected to be earlier than that of discordant pairs. Obtaining a precise estimate of the within-person relationship of age at onset with disorder liability is more difficult than gaining an eyeball impression. The approach we take provides a maximum likelihood estimate of the association. It also has the valuable feature of correcting for censoring the estimates of relatives’ correlations for disorder liability.

### Pairs of Relatives

If both relatives are affected, their joint likelihood can be regarded as a single region of the four-variate multivariate normal distribution. Both members of the pair must be above the liability threshold, and their ages at assessment (Age_1_ and Age_2_) must be greater than their respective ages at onset (AO_1_ and AO_2_. Using subscripts 1 and 2 for relative 1 and relative 2, the joint likelihood is therefore:

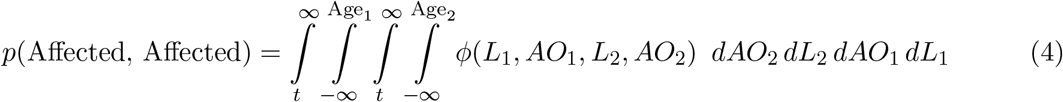

This formulation is appropriate when only affection status and age at assessment are known. If ages at onset are known, the likelihood reduces to a double integral of the four-dimensional multivariate normal distribution, as follows:

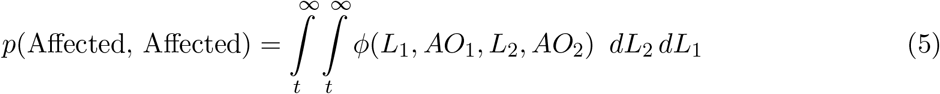

The likelihoods for discordant pairs can be described as the sum of two components, similar to Equation 3:

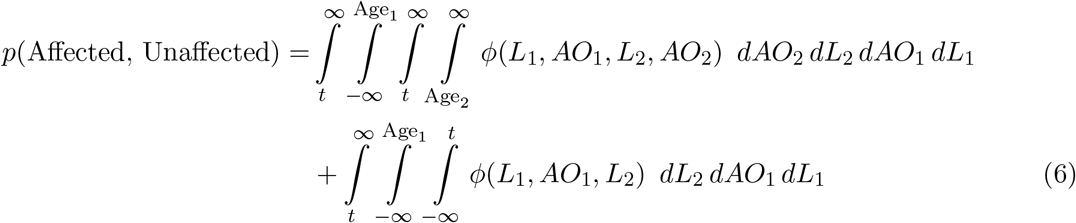

Again in the event that actual age at onset is known, it is no longer necessary to integrate over that dimension. As a result, the likelihood simplifies to:

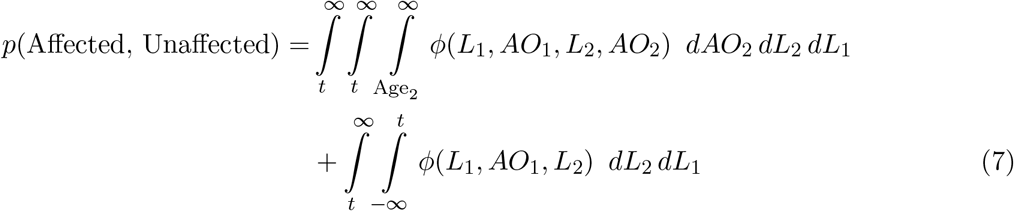

The likelihood for an Unaffected-Affected pair is equivalent except for exchanging the subscripts 1 and 2:

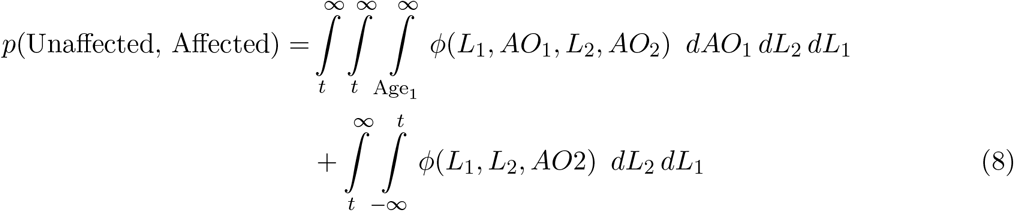

There are four possible ways for concordant unaffected pairs to be observed. One is where both are below the liability threshold. The second and third are where one relative is below the liability threshold, and the other is above the threshold but younger than their age at onset. The fourth is where both are above the liability threshold, but both are younger than their ages at onset. In mathematical terms we have:

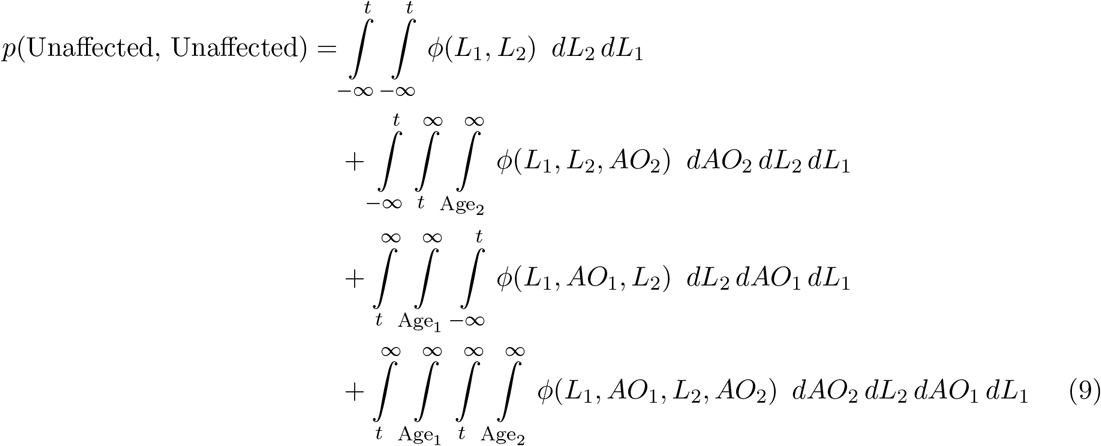

This approach corrects the parameter estimates for censoring, to yield asymptotically unbiased maximum likelihood estimates of the parameters in the path diagram shown in Figure 2.

**Figure 2.**
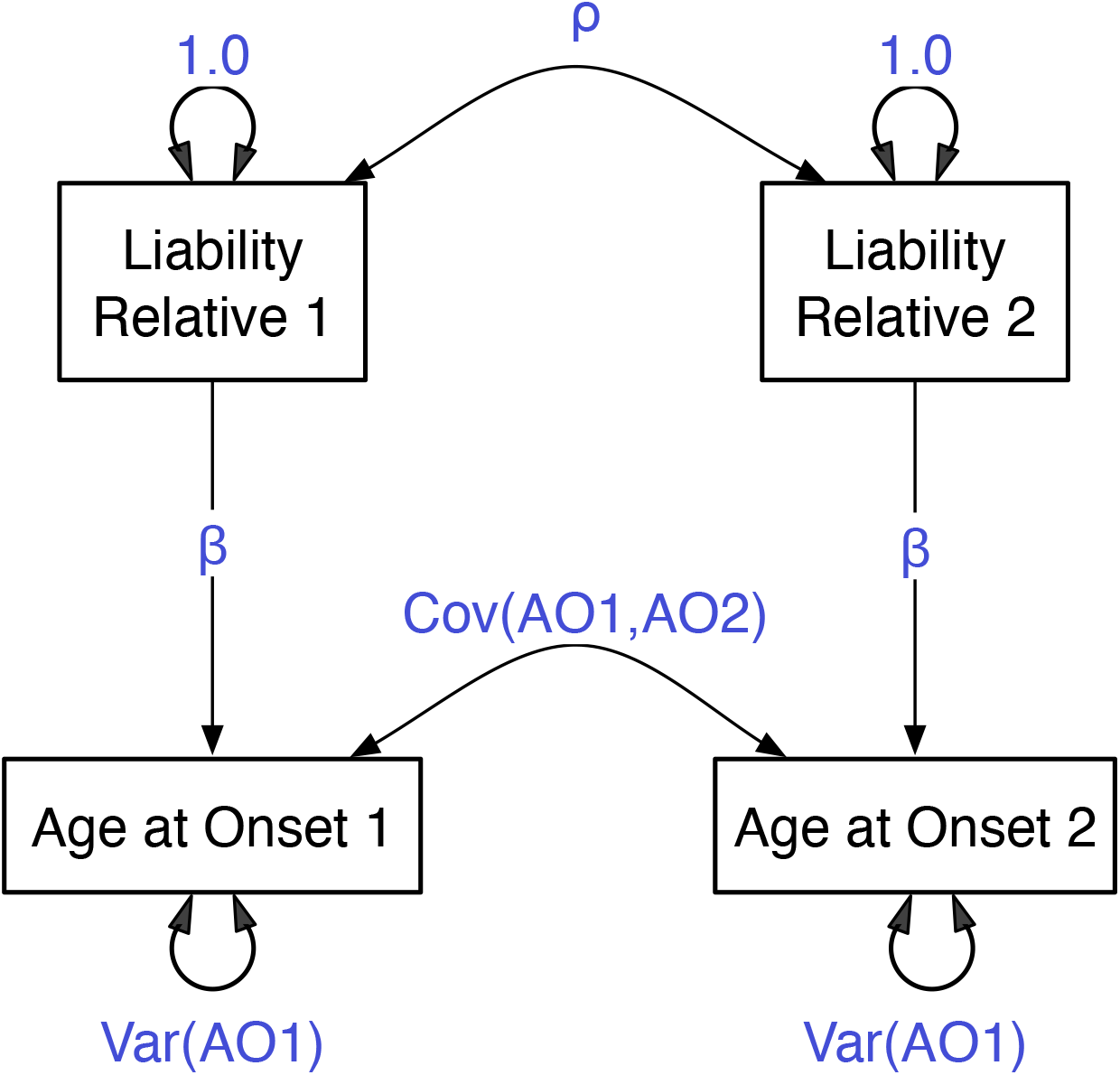
Path diagram of age at onset and liability model for pairs of relatives. The Var(AO1) and Cov(AO1,AO2) parameters are respectively measures of the residual variation in, and covariation between, relatives’ ages at onset

### Implementation

#### General considerations

To fit such a model to data from pairs of relatives, one could write a standalone program. Doing so would have several disadvantages compared to using an existing package that can estimate parameters using full information maximum likelihood (FIML). The disadvantages include: additional software maintenance, difficulty in extending it to handle extensions such as covariates, and the duplication of effort to handle the input data and to format output. Although the R package OpenMx Neale et al., 2016 was not designed for such analyses, it has the necessary FIML estimation, which can be used for both continuous and ordinal traits, or a combination thereof Pritikin et al., 2018. Additional features that make the approach feasible include that it can model mixture distributions and make use of definition variables, each of which will be described below. We now consider how this functionality can be applied to the age-at-onset case.

#### Mixture Distribution

While OpenMx’s FIML can directly evaluate the joint likelihood of continuous and ordinal variables, the two different ways of being unaffected (being below the liability threshold or above it, but with onset at a later date than the assessment) complicate matters. All possible scenarios by which the data could have arisen, and their likelihoods, must be considered. Here, ambiguities concerning whether an individual is unaffected due to being below threshold on the liability dimension, or unaffected because they have not reached their age at onset generate alternative scenarios, which are represented as the summed parts of expressions such as Equation 9.

The summation of multiple components of likelihood expressions, such as Equation 9, is not commonly encountered when specifying structural equation models. One way to evaluate such likelihoods is to treat them as a mixture distribution with unit weights for the different components. Finite mixture distributions are often used to handle situations where observations might be of more than one type. An example is the analysis of data from twin pairs whose zygosity is not known Neale, 2003. In general, the likelihoods of mixture distributions can be evaluated as:

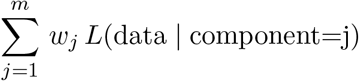

where *m* is the number of components, *w*_*j*_ is the weight of the component *j*, and *L*(*data*|*component* = *j*) is the likelihood of the data under the model for component *j*. In latent class analysis, the weights are estimated as the proportions of the population that belong to each class, which causes some statistical problems in model comparison. By contrast, here the weights are not estimated, merely fixed at 1 to sum the likelihood components. Figure 3 shows a schematic diagram of the OpenMx script structure.

**Figure 3.**
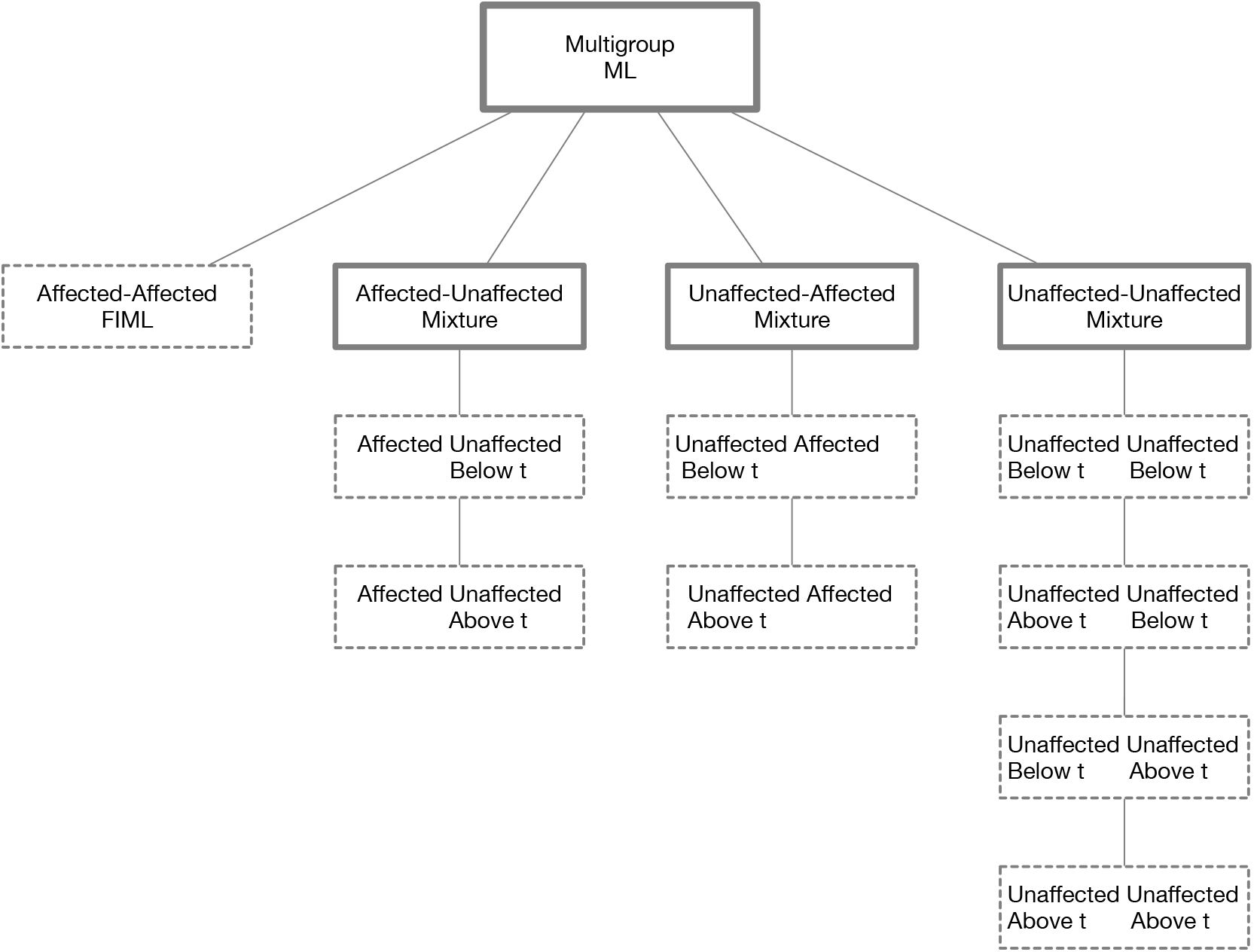
Schematic of multigroup mixture distribution model for age at onset and disorder. Boxes with dotted lines represent mxModels evaluated with joint continuous-ordinal FIML. The top level mxModel aggregates the components from the four group types. The Mixture mxModels sum the likelihoods from their component submodels.

### Multiple data vectors

Usually, when fitting models to mixture distributions, a single row of data is used to evaluate the likelihood of each of the components. The present situation is more difficult because *different data vectors are needed to evaluate the different components of the likelihood*. Data preparation therefore involves generating additional data vectors to represent the alternative states of being unaffected. Two vectors are required for discordant pairs, and four for the concordant unaffected pairs.

#### Concordant affected pairs

Let the data vector for pairs of relatives contain three variables per person: their age at last assessment, their affection status, and their age at first onset if they are affected. Affection status is a binary variable, while age at onset can be considered to be continuously distributed. The likelihood of a concordant affected pair, shown in Equation 5, is straightforward to evaluate as it has only one term, which integrates over two of the four-dimensional normal distribution of the pair’s liability dimensions and their observed ages at onset. This likelihood can be evaluated by providing OpenMx with a data vector such as:

|  |  |  |  |  |  |  |  |
| --- | --- | --- | --- | --- | --- | --- | --- |
| *Age1* | *Age2* | *L1* | *AO1* | *AO1b* | *L2* | *AO2* | *AO2b* |
| 28 | 29 | 1 | 25.5 | NA | 1 | 24 | NA |

When a person’s age at onset is known, it can be treated as a continuous variable in a joint continuous and ordinal FIML analysis. However, when someone is unaffected, if we assume that they are above threshold, their age at onset must be in the future. Therefore, the lower limit of their integral over the age-at-onset distribution should be their last age at assessment, and the upper limit can be set to infinity. Thus, we see that in some cases the age-at-onset variable is a binary variable, while in others it is observed and continuous. Our approach to handling this situation is to make data vectors that contain columns for both the binary and continuous representations of the data, as tabled above.

#### Discordant affected-unaffected pairs

A typical data vector for discordant pair might be appear as follows:

|  |  |  |  |  |  |  |  |
| --- | --- | --- | --- | --- | --- | --- | --- |
| *Age1* | *Age2* | *L1* | *AO1* | *AO1b* | *L2* | *AO2* | *AO2b* |
| 25.5 | 23 | 1 | 25.5 | NA | 1 | NA | 1 |

Here, affected relative 1 is above threshold (L1=1), their observed age at onset is 25.5 years (AO1=25.5), their binary variable for onset is missing (AO1b=NA). Unaffected relative 2 is also above the liability threshold (L2 = 1), but has not passed their age at onset (AO2=NA and AO2b=1). This data vector evaluates only the first term in Equation 6.

The second term in the likelihood of a discordant affected-unaffected pair (Equation 6) can be evaluated with the following data vector:

|  |  |  |  |  |  |  |  |
| --- | --- | --- | --- | --- | --- | --- | --- |
| *Age1* | *Age2* | *L1* | *AO1* | *AO1b* | *L2* | *AO2* | *AO2b* |
| 25.5 | 26 | 1 | 25.5 | NA | 0 | NA | NA |

To add this likelihood to that of the first term, we employ a two-part mixture distribution, with equal unit weights.

Although the data vectors being evaluated differ according to the pair’s affection status, the expected covariances, means, and thresholds are constrained equal across all components. Age1 and Age2, the last dates of assessment of the pair of relatives, are used as definition variables (Boker et al., 2011; Eaves et al., 1996; Neale, 1998) to define the lower limit of integration for those not yet at their age at onset. They do not appear in the model’s expected covariances, means, or thresholds.

The variable pairs AO1 with AO1b, and AO2 with AO2b are different measures of exactly the same variable. AO1 is continuously measured, while AO1b is a binary measure of the same distribution. Therefore, these two variables have the same variances and they covary this amount to represent their perfect correlation. Similarly, their means are identical. The structure of the covariance matrix of the vector of variables has the following form:

|  | *L1* | *AO1* | *AO1b* | *L2* | *AO2* | *AO2b* |
| --- | --- | --- | --- | --- | --- | --- |
| *L1* | 1 | *cLA* | *cLA* | *crLL* | *crLA* | *crLA* |
| *AO1* | *cLA* | *vAO* | *vAO* | *crLA* | *crAA* | *crAA* |
| *AO1b* | *cLA* | *vAO* | *vAO* | *crLA* | *crAA* | *crAA* |
| *L2* | *crLL* | *crLA* | *crLA* | 1 | *cLA* | *cLA* |
| *AO2* | *crLA* | *crAA* | *crAA* | *cLA* | *vAO* | *vAO* |
| *AO2b* | *crLA* | *crAA* | *crAA* | *cLA* | *vAO* | *vAO* |

The variance of the disorder liability dimensions *L*1 and *L*2 are set to 1.0 as they are latent variables. The variance of the age-at-onset variable *vAO* appears eight times in the matrix, covering the variances and within-person covariances for the continuous and binary instances of these variables. Within-person association between liability to the disorder and their age at onset is estimated with parameter *cLA*. Across-relative correlations are in the 3×3 off-diagonal blocks and denoted *cr*. They estimate the covariances across relatives for disorder liability (*crLL*), for age at onset (*crAA*) and across relatives across dimensions (*crLA*). The vectors for estimating means and thresholds are similarly structured, with zero means for the disorder liabilities but the same free parameter for the mean of the four age-at-onset variables. The only threshold to be estimated is that of the disorder liability dimension, which is set equal across the two relatives and across all components of likelihood for all four types of pair.

A critical part of this data preparation is that no individual should have values for *both* AO1 and AO1b: one of the two must always be missing, coded as NA. This is essential because the expected covariance between these variables is the same as their variance, since they are identical variables but observed in one of two forms. That is, the age at onset is either continuous, for a known age in the past, or above current age, and represented as a binary variable for some unknown time in the future. To evaluate the likelihood taking into account the person’s last age at assessment, the lower limit of the integral is set using the participant’s age as a definition variable in OpenMx.

#### Concordant unaffected-unaffected pairs

The expression for the likelihood for pairs of this type contains four terms, per Equation 9. Accordingly, four data vectors are generated to evaluate them, which are summed in a four-part mixture distribution. The data vectors needed for the four terms are:

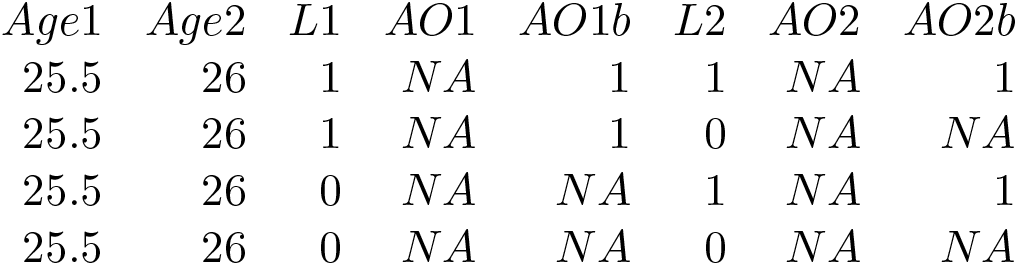

| *Age1* | *Age2* | *L1* | *AO1* | *AO1b* | *L2* | *AO2* | *AO2b* |
| --- | --- | --- | --- | --- | --- | --- | --- |
| 25.5 | 26 | 1 | NA | 1 | 1 | NA | 1 |
| 25.5 | 26 | 1 | NA | 1 | 0 | NA | NA |
| 25.5 | 26 | 0 | NA | NA | 1 | NA | 1 |
| 25.5 | 26 | 0 | NA | NA | 0 | NA | NA |

Software to generate the necessary vectors from the original is written in R, along with the OpenMx model specification and data simulation. The user only has to prepare data files with six columns: the relatives’ last age at assessment, their affection status, and their age at onset if observed. This resource can be found in the supplementary materials.

### Simulation

We tested the ability of the model-fitting approach to recover population parameter estimates via simulation. The simulated data consisted of six variables, three for each relative: Age at Last Assessment (continuous), Affection Status (binary 0/1 for below/above), and Age at Onset (continuous or NA if unaffected). The data were simulated according to the covariance matrix, means and thresholds shown in Table 1. Ages of the relatives were also simulated from a normal distribution, although other distributions would be feasible. The data were then processed to replace Affected (1) with Unaffected (0) in all cases whose age at onset was later than their age at the last assessment. Covariances between the age distributions of the relatives were set at .8, and the age distributions were uncorrelated with the age at onset and liability variables, yielding the covariance matrix in Table 1 The consequences of censoring can be seen in the scatterplot of a sample of the data, shown in Figure 5.

**Table 1.** Covariance matrix for simulation. Means were set to zero, and the threshold for the liability distribution was set to the 40^th^ percentile.

|  | Age1 | Age2 | Liability1 | Age at Onset1 | Liability2 | Age at Onset2 |
| --- | --- | --- | --- | --- | --- | --- |
| Age1 | 1 | 0.8 | 0 | 0 | 0 | 0 |
| Age2 | 0.8 | 1 | 0 | 0 | 0 | 0 |
| Liability1 | 0 | 0 | 1 | 0.7 | 0.6 | 0.4 |
| Liability2 | 0 | 0 | 0.7 | 1 | 0.4 | 0.6 |
| Age at Onset1 | 0 | 0 | 0.6 | 0.4 | 1 | 0.5 |
| Age at Onset2 | 0 | 0 | 0.4 | 0.6 | 0.5 | 1 |

These data were then further processed with a function `assign_affection_data()` to generate the additional data vectors to evaluate the multiple terms of the likelihoods for the pairs that include at least one unaffected relative (Equations 7 to 9).

The right panel of Table 2 shows the estimates recovered when the sample size was set to 100,000 pairs, and there is close agreement with the values used to generate the data.

**Table 2.** Recovery of population values from fitting the censored model for age at onset. Sample size was set to 100,000 pairs.

|  | Simulated Correlations |  |  |  | Parameter Estimates |  |  |  |
| --- | --- | --- | --- | --- | --- | --- | --- | --- |
|  | L1 | L2 | AO1 | AO2 | L1 | L2 | AO1 | AO2 |
| L1 | 1 | 0.7 | 0.6 | 0.4 | 1.000 | 0.693 | 0.580 | 0.385 |
| L2 | 0.7 | 1 | 0.4 | 0.6 | 0.693 | 1.000 | 0.385 | 0.580 |
| AO1 | 0.6 | 0.4 | 1 | 0.5 | 0.580 | 0.385 | 1.000 | 0.488 |
| AO2 | 0.4 | 0.6 | 0.5 | 1 | 0.385 | 0.580 | 0.488 | 1.000 |

## Results

Figure 4 shows a comparison between estimating the correlation in liability in the age-of-onset model and simply estimating the tetrachoric correlation between the observed affection statuses of the pairs of relatives. For this particular amount of censoring, the two estimates diverge: at higher population correlations (over .4 in this example) the tetrachoric is biased downwards, whereas for true population values less than .4 there is an upwards bias. This result diverges from those observed when the distribution is truncated due to factors other than age at onset (Neale, Eaves, Kendler, & Hewitt, 1989). The pattern observed here suggests that estimates of correlations between relatives, when some of the sample are still at risk of having the disorder at a later age, will tend to underestimate the heritability in studies of MZ and DZ twins. Overestimation of the effects of common environment factors are also expected as a result of censoring.

**Figure 4.**
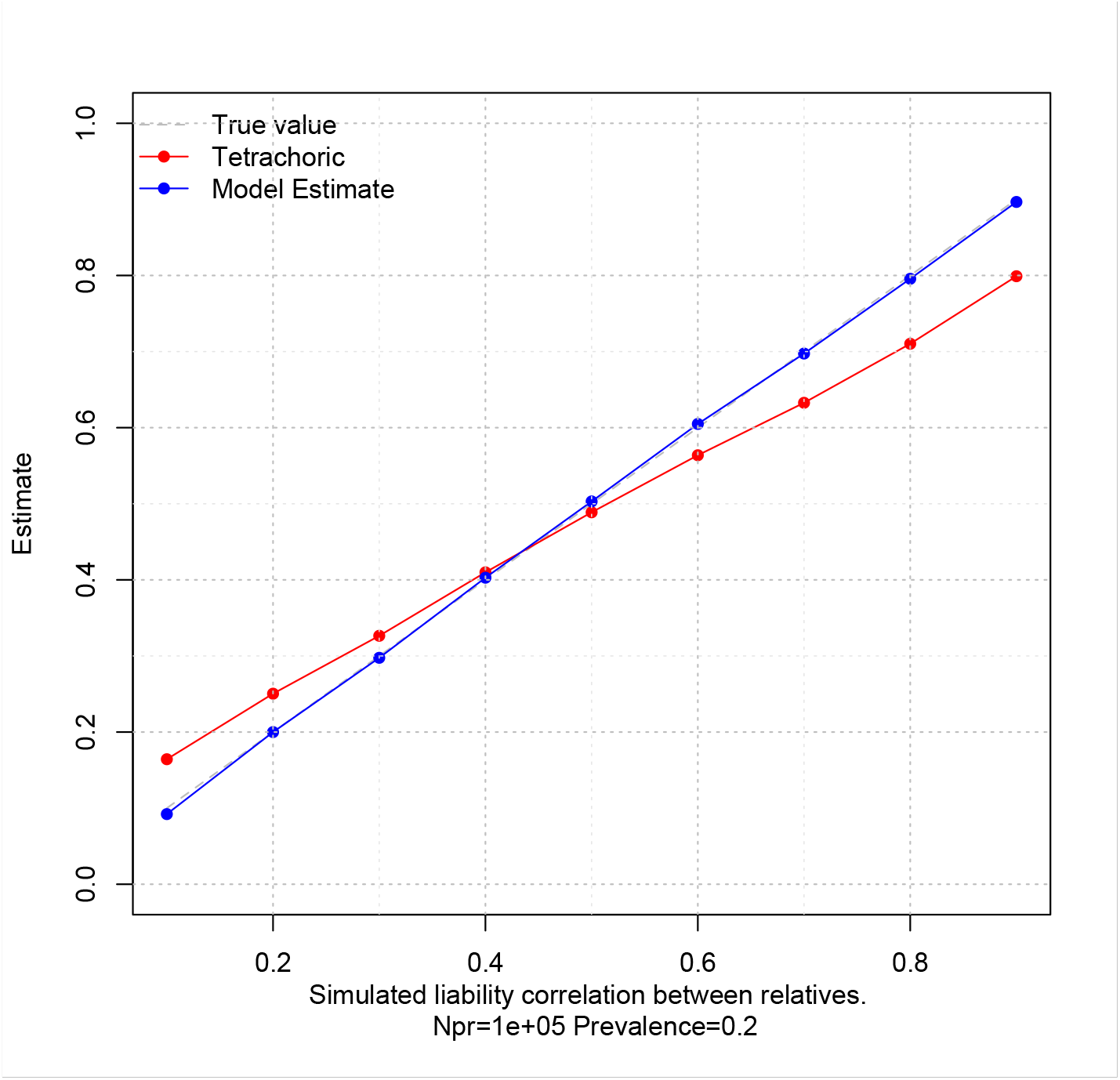
Difference between model-recovered estimate of correlation in liability from a censored population and tetrachoric correlation of the observed affection status

**Figure 5.**
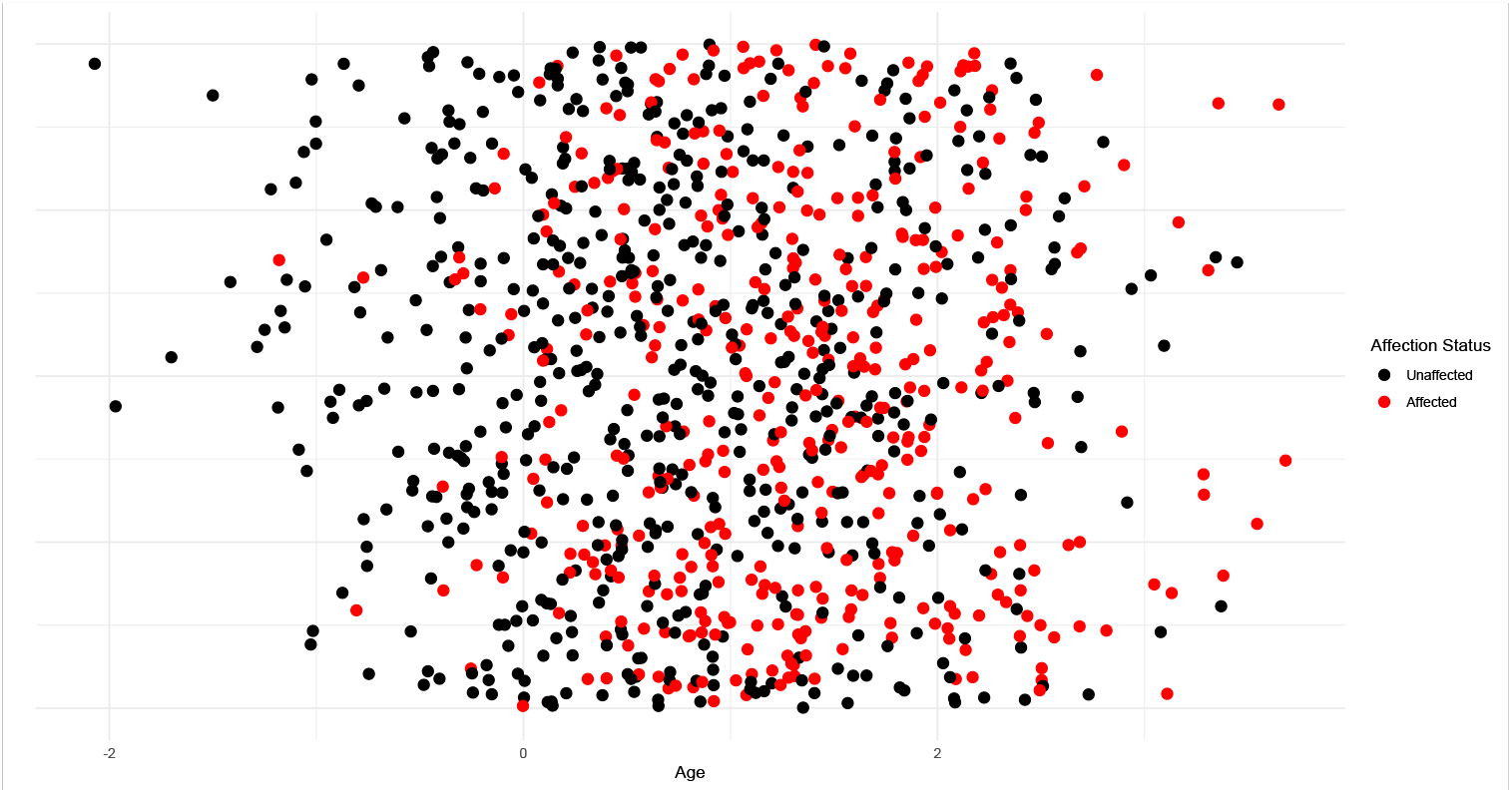
Scatterplot of simulated data showing that the proportion of affected individuals (red dots) increases with age. Y-axis values were jittered.

### Application

To illustrate, we fit the age-at-onset model to data from 756 MZ twin pairs (327 male, 427 female) on cannabis use and age-at-onset in the Adolescent Behavior Development Study (ABD), which collected data from young twins and their parents in Virginia in the early 1990s Eaves et al., 1997; Hewitt et al., 1997. The maximum likelihood estimates and 95% confidence intervals Neale and Miller, 1997 of the model parameters are shown in Table 3. The key estimate of −.21 for the causal path from liability to cannabis use to the age at onset of use is not standardized because the variance of age at onset does not equal unity. Following standardization, the estimate is .212/sqrt(.097) = −0.682, which is consistent with almost half of the variance in age at onset being associated with the liability to initiate. The estimated mean age at onset of 18.12 years for the sample is higher than the mean ages at onset of the concordant and discordant pairs, which are 16.8 and 17.8 years, respectively. Although it may seem odd that the estimated mean is higher than the observed mean of onset, this is a natural consequence of part of the sample being still at risk for initiation. Similarly, the estimate of the liability threshold of z = −.315 yields 62.4% of individuals who are above the threshold, which is greater than 48.5% of individuals who have already started use, suggesting that approximately 14% are expected to onset at a later age.

**Table 3.** Parameter estimates with 95% confidence intervals Neale and Miller, 1997 for the model shown in Figure 2.

| Parameter | Lower 95% | Estimate | Upper 95% |
| --- | --- | --- | --- |
| $\beta$ | -0.263 | -0.212 | -0.152 |
| Var(AO1) | 0.042 | 0.052 | 0.063 |
| Cov(AO1,AO2) | 0.030 | 0.039 | 0.049 |
| Mean Age at Onset | 17.68 | 18.12 | 18.55 |
| Liability threshold | -0.315 | -0.224 | -0.134 |

## Discussion

This article presents a method for measuring the covariance between age at onset and liability to a disorder. This information is not available when only data from unrelated persons are available. The method has the considerable advantage of controlling for censoring due to individuals not having reached their age at onset, which to our knowledge has not been successfully implemented before. Our prior work on control for censoring through contingency table analysis (Pickles et al., 1994) involved treating ages as ordinal data, which limited the precision of the method. In addition, the present treatment explicitly differentiates between liability to disorder and liability to age at onset, and estimates this correlation both within and across relatives.

The application to data on cannabis initiation revealed a moderately high correlation of −.68 between onset liability and disorder liability. This result is consistent with higher liability among those with earlier onset. Risk factors for use – such as polygenic scores – would be expected to be higher among those with earlier onset. Conversely, polygenic scores for early onset would likely be higher among affected than unaffected individuals. The addition of covariates such as polygenic scores and demographic variables to the model is a relatively straightforward extension that we plan to implement, but it is one of many possibilities.

First, it is relatively straightforward to analyze data from multiple types of relatives, by adding groups that are structured the same way as Figure 3. Thus a classical twin study could be modeled in the usual way (Verhulst et al., 2019). With large central databases of medical records, combined with information on who is related to whom (e.g., those in Denmark) both large sample sizes and a variety of types of relatives could be constructed. In principle, no individual should contribute to more than one pair of relatives, since such observations are nonindependent. Ideally, one would extend the method to handle pedigree data, but this seems difficult to do beyond very small pedigrees. The number of mixture components for a pedigree of size *N* would be 2^*N*^, e.g., 1024 for a pedigree of size 10. The computational time would also grow exponentially and numerical integration precision could become an issue. If a correction for the standard errors could be found, then using multiple relative types with some overlapping individuals would be an attractive practical approach. In large databases, it may be practical to ensure that each individual appears only once in the data, while retaining high statistical power for the analysis.

A second set of extensions concerns the multivariate case, although expanding beyond two disorders seems difficult. In the bivariate case, there would be 16 components to the likelihood of pairs concordant for having neither outcome. The possible pair types would also increase from 4 to 16, further increasing the computational burden but still within practical limits. Extensions beyond the bivariate case seem practical for traits or disorders for which the population has already passed the age at risk of onset. These additional variables could be measured at the binary, ordinal, or continuous level. Continuous variables would increase the number of variables in the integrands of Equations 7 to 9, but would not change number of dimensions over which integration would occur. There would be a limit to the number of binary or ordinal variables that could be added, since numerical integration of the multivariate normal distribution becomes exponentially more difficult with more dimensions - known as the curse of dimensionality. A full bivariate model that estimates all the covariances between the disorder and onset variables both within and between persons would be a natural place to start. In some cases, simply adding covariates whose fixed effects are to be regressed out would be the limit of the multivariate analysis. Within computational limits, this approach seems practical and of considerable value, since understanding the factors that influence age at onset, and through what pathways they do so could potentially alleviate the burden of disease for the population. Later onset naturally incurs lower health care costs, and can do so completely if age at onset is delayed beyond the lifespan.

Although this article has focused on models for diagnostic outcomes, we should recognize that their applicability may be much broader. Most measures of psychological states, psychiatric disorders and substance use begin at the item level, often with a factor model for assessing the latent trait of interest. In many cases, these items will also have variable ages at onset. Substance use items clearly show variable age at onset, as do many items contributing to psychopathology, cognitive ability and other traits. Another area of likely valuable applications is the study of mild cognitive impairment or dementia, where censoring is likely to be encountered due to withdrawal from the study or death. Correcting for censoring seems important in many contexts, but perhaps especially so in studies of relatives.

## Data Availability

All data produced in the present study are available upon reasonable request to the authors

https://vipbg.vcu.edu/vipbg/Articles/

## 1 Acknowledgments

MCN was supported by NIH grant U01 DA051037.

